# A cross-sectional questionnaire-based landscaping of female infertility reveals genital Infections as a major contributor to reproductive tract anomalies, menstrual disorders, and infertility

**DOI:** 10.1101/2023.09.04.23295020

**Authors:** Naresh Patnaik, Uttam Sarkar, Malathi Jojula, Hema Vaddiraju, Ruchi Jain Dey

## Abstract

**Objectives:** Female infertility is a global health concern. The association of genital infections with female infertility is neglected due to their chronic but asymptomatic nature. Lack of routine diagnosis and delay in treatment further causes intractable pathological sequalae and consequential infertility. This study aims to identify the most significant prognostic symptoms of genital infection(s) that correlate strongly with reproductive tract anomalies, menstrual disorders, and infertility.

**Methods:** We designed a detailed questionnaire and conducted a cross-sectional study with 100 female subjects, categorized into infertile (n1 = 62) and healthy groups (n2 = 38). The data collected was documented and statistically analyzed.

**Results:** This study highlights an early onset of infertility (21-30 years). Almost 27% of the infertile subjects are symptomatic for genital infections and ∼42% exhibit menstrual irregularities. Polycystic ovarian syndrome/disease (PCOS/PCOD, ∼30%), are observed to be the most predominant disorders followed by endometrial disorders (∼10%) and tubal damage (∼8%) in infertile subjects. A multivariate correlation analysis revealed a highly significant (*p* ≤ 0.05) and strong association (0.15 < Φ ≤ 1.0) between menstrual disorders, endometrial disorders, uterine/tubal blockage, and hormonal disruption with infection-associated symptoms, such as vaginitis, cervicitis, pelvic inflammatory disorder (PID), dyspareunia, tuberculosis (TB), urinary tract infection (UTI), sperm, and semen abnormalities.

**Conclusions:** Our study reveals genital infections to be a significant contributor to female infertility. The questionnaire designed here offers a useful tool for self or clinical assessment and may help in timely prognosis/diagnosis of genital infections which may contribute to improved management of reproductive health and fertility.

**Synopsis:** The study reveals impact of genital infections on female infertility and offers a comprehensive questionnaire-based tool for an early self/clinical prognosis of infection induced infertility.

## Introduction

Infertility and menstrual disorders are some of the major health concerns affecting women worldwide. A recent report by WHO estimates, that approximately 1 in 6 women (17.5% of the adult population) are affected by infertility [1–3], and the rates of infertility are comparable across low-, middle- and high-income countries [3]. The significant number of individuals impacted by infertility, menstrual disorders, and reproductive tract anomalies highlights the necessity to broaden the availability of reproductive health care and prioritization of this matter in health research and policy [3]. This will ensure the availability of secure, efficient, and reasonably priced treatment modalities for women suffering from these disorders and couples desiring to become parents [2, 3].

Infertility is a disease of both the male and/or female reproductive system and is defined as the inability to conceive after a year of regular, unprotected sexual intercourse [2, 3]. It is a complex and multi-factorial disorder that can result from numerous factors such as genetic, environment, nutrition, infections, and lifestyle [4]. In the absence of regular reproductive health assessment due to cultural/financial constraints and societal stigma, reproductive health issues remain largely ignored. This leads to delayed diagnosis and treatment of these disorders causing infertility in the long run [2]. Infertility has the potential to cause substantial emotional and economic strain on the families and impacts individuals’ mental and psychosocial welfare. Countless individuals encounter overwhelming medical expenses during infertility treatments, leading to healthcare-induced poverty [2, 5–7].

In tropical countries like India, where tuberculosis (TB) and other infectious diseases are widespread, the role of infectious diseases in causing infertility becomes a prominent consideration as indicated by several studies [5, 8–10]. Genital infections (sexually transmitted, postpartum, post-abortion infections and secondary infections e.g., TB) may cause a wide spectrum of conditions like vaginitis, cervicitis, dyspareunia and PID [11, 12], endometriosis and other endometrial disorders [5, 13], PCOS/PCOD, ovarian atresia or cysts and anovulation [14, 15]. These infections are generally chronic in nature and cause inflammation and irreversible scarring of the reproductive tract leading to sub-fertility, infertility, recurrent abortions, low birth weight, pre-term birth, stillbirth, maternal and infant mortality [5, 9, 10]. For instance, female genital tuberculosis (FGTB) is estimated to be responsible for 6-25% of female infertility cases in India [5, 16–19]. FGTB can occur as a primary infection or as a result of secondary spread from infected lungs or reactivation of the latent infection and may cause reduced ovarian reserve (ovarian atresia), PCOS/PCOD, fallopian tube, and uterine damage (e.g. adhesions, cysts and endometrial disorders), pregnancy failures and infertility. The treatment of FGTB involves a combination of anti-TB drugs and surgical intervention(s), however, the success rates of these treatments in terms of the improvement in fertility vary depending on the severity of the reproductive tract damage [20–22]. Previous research also links bacterial vaginosis (BV) to PID and endometritis with consequential infertility [11]. Endometrial presence of BV-related bacteria is connected to 3.4 times higher infertility risk [13]. Women with subclinical PID show 40% lower pregnancy chances compared to those without it [12].

Substantial clinical evidence exists for the role of infections in infertility from investigational surveys involving confirmatory tests. However, questionnaire-based surveys for female infertility do not typically include queries regarding infection-associated symptoms which can be self/clinically assessed. Hence, to understand the landscape of infertility and the influence of infections on various reproductive health parameters among female subjects in the rural setup, we designed a detailed questionnaire that specifically addresses the gaps listed above by the inclusion of detailed queries on infection-associated symptoms. We then conducted an observational, cross-sectional survey at an OB & GYN hospital (mother and child care hospital) in Warangal, India. The subjects included were mainly from rural and semi-urban backgrounds. Our study provides evidence for usefulness of the questionnaire designed by us as a valuable tool for self or clinical assessment of female reproductive health and may help in timely prognosis/diagnosis of genital infections which may further improve the management of reproductive health and fertility.

## Materials and Methods

### Ethics statement

The research was conducted in accordance with the ethical guidelines set forth by the Indian Council of Medical Research (ICMR) for biomedical and health research involving human participants [23]. The study was approved by the Institutional Human Ethics Committee (IHEC) of Birla Institute of Technology and Sciences (BITS) Pilani Hyderabad campus (Protocol number: BITS-HYD/IHEC/2022/01). All individuals who participated in the study were fully informed about various aspects, including the study’s objectives, methodologies, sources of funding, potential conflicts of interest, institutional affiliations of the researcher, anticipated benefits, potential risks, and the discomfort they might experience. Their participation was contingent on obtaining their informed consent. Furthermore, to safeguard patient confidentiality, we took measures to de-identify patient information. This included the removal of exact ages, which were replaced with age ranges, and the omission of exact dates or photographs during presentation of the data (See **Supplementary Material 1** for patient consent form).

### Study design, inclusion/exclusion criteria, and enrolment

The observational, cross-sectional survey was conducted in Curewell Hospital, Warangal, Telangana, India. Subject recruitment for the study was done based on the gynecologist’s assessment. Subjects were enrolled in the study based on the inclusion/exclusion (I/E) criteria. In this study, female subjects (18-45 years) visiting the clinic for infertility treatment (along with their male partners) were enrolled. Female subjects outside the age group of 18-45 years were excluded from this study. Subjects (couples) with complaints of infertility (not getting conceived for a duration of ≥12 months) were classified into the infertile group. Infertile subjects were further stratified into primary (no live birth) and secondary infertility cases (at least one live birth). Females with regular menstrual cycles and no recent history of infertility or genital infection were classified into the healthy control group. Subjects diagnosed with Human Papillomavirus (HPV), Human Immunodeficiency virus (HIV), and Hepatitis C virus (HCV), and/or undergoing any antibiotic or hormonal therapy or *in vitro* fertilization (IVF) treatment, were excluded from this study.

### Study Population and questionnaire-based data collection

The sample size (n = 97) was determined by the help of formula n=[DEFF * Np(1-p)]/[(d^2^/Z^2^_1-α/2_*(N-1) +p(1-p)], where n = sample size, DEFF = design effect, N= population size (10^7^), p = estimated proportion (6.7% ± 5), q= 1-p, d = desired absolute precision (5%) or absolute level of precision, achieving a confidence level of 95% [24, 25]. Hence, a total of 100 female subjects were enrolled for this study based on I/E criteria defined above following the informed consent. These subjects were further allocated into groups (infertile and healthy) based on their fertility status (**Figure 1** for study outline). The subjects in each group were assessed on a single time-point using a detailed questionnaire. The questionnaire (**Supplementary Material 2**) is designed to collect a comprehensive data on the female subjects included a variety of parameters (**Supplementary Material 3, Table S1).** The study solely assessed the existing information provided by the subjects (including previous test reports) and did not involve collection of any new samples or performance of any new tests.

**Figure 1.**
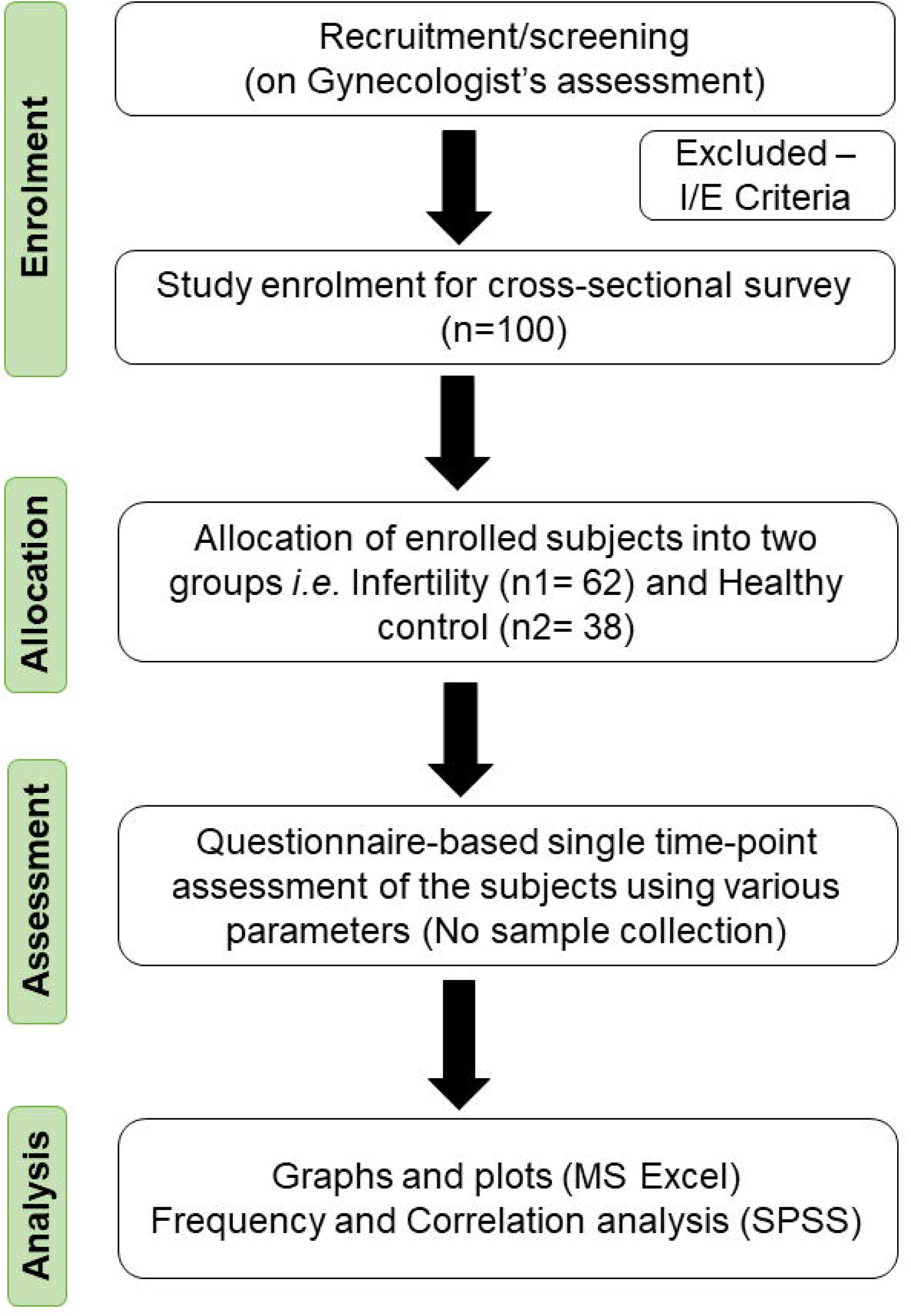
Study design flowchart. The figure depicts an outline of the cross-sectional survey conducted on infertile and healthy women to understand various parameters influencing female fertility. In this study, 45 parameters (listed in Table S1) were assessed to determine the frequency distribution and Phi correlation between different parameters (as described in Table S2A-D and Figure 9, respectively).

### Data handling and statistical analysis

The data was collected through questionnaire followed by digital recording into Microsoft (MS) Excel sheet via Google forms. For the purpose of analysis, the data sets were simplified into a multinomial format (Yes/No/No data). Data was plotted using MS Excel. Data analysis for calculation of the frequency distribution and correlation (Phi coefficient) was performed using IBM SPSS Statistics for Windows, Version 25.0., IBM Corp., Chicago, ILSPSS amongst multiple parameters listed in **Table S1**. Data is reported as per the STrengthening the Reporting of OBservational studies in Epidemiology (STROBE) guidelines (https://www.strobe-statement.org/).

## Results

### Landscape of female Infertility and other reproductive health-related conditions

A comprehensive questionnaire-based cross-sectional study is carried out to identify the most significant parameters associated with subfertility, infertility (primary and secondary), or conditions like pre-term birth, spontaneous abortions, medical termination of pregnancy, and ectopic pregnancy in females. Out of the 100 subjects assessed, 62 were categorized into the infertile group and 38 were healthy subjects with no history of infertility or menstrual disorders (**Figure 1**). Amongst the infertile group, only female infertility (39%) with healthy and fertile male spouses, predominated the assessed population (**Figure 2A**). Infertility due to only male factors with apparently healthy/fertile females represented only 2% of the study population (**Figure 2A**). Female infertility is seen in 60% of the total study population including 21% of cases contributed by both male and female infertility (**Figure 2A**). Out of the total 62 infertility cases, 90% are diagnosed with primary infertility, rest 10% are cases of secondary infertility (**Figure 2B**). Our study highlights an early onset of female infertility as young as ≤ 20 years of age **(Figure 2C)**, with predominance of both primary and secondary infertility in the age group of 21-30 years **(Figure 2C)**. Among 62 infertility cases, ∼74% failed to conceive even once, and a large proportion of ∼23% suffered from spontaneous abortions/have undergone medical termination of pregnancy (MTP) (**Figure 2D**). Ectopic pregnancies were also observed in ∼3% of cases, however, no pre-term birth cases were observed in this study population (**Figure 2D**). Age distribution of these failed pregnancies, reveals an early occurrence in the age group of 21-25 years (**Figure 2E**).

**Figure 2.**
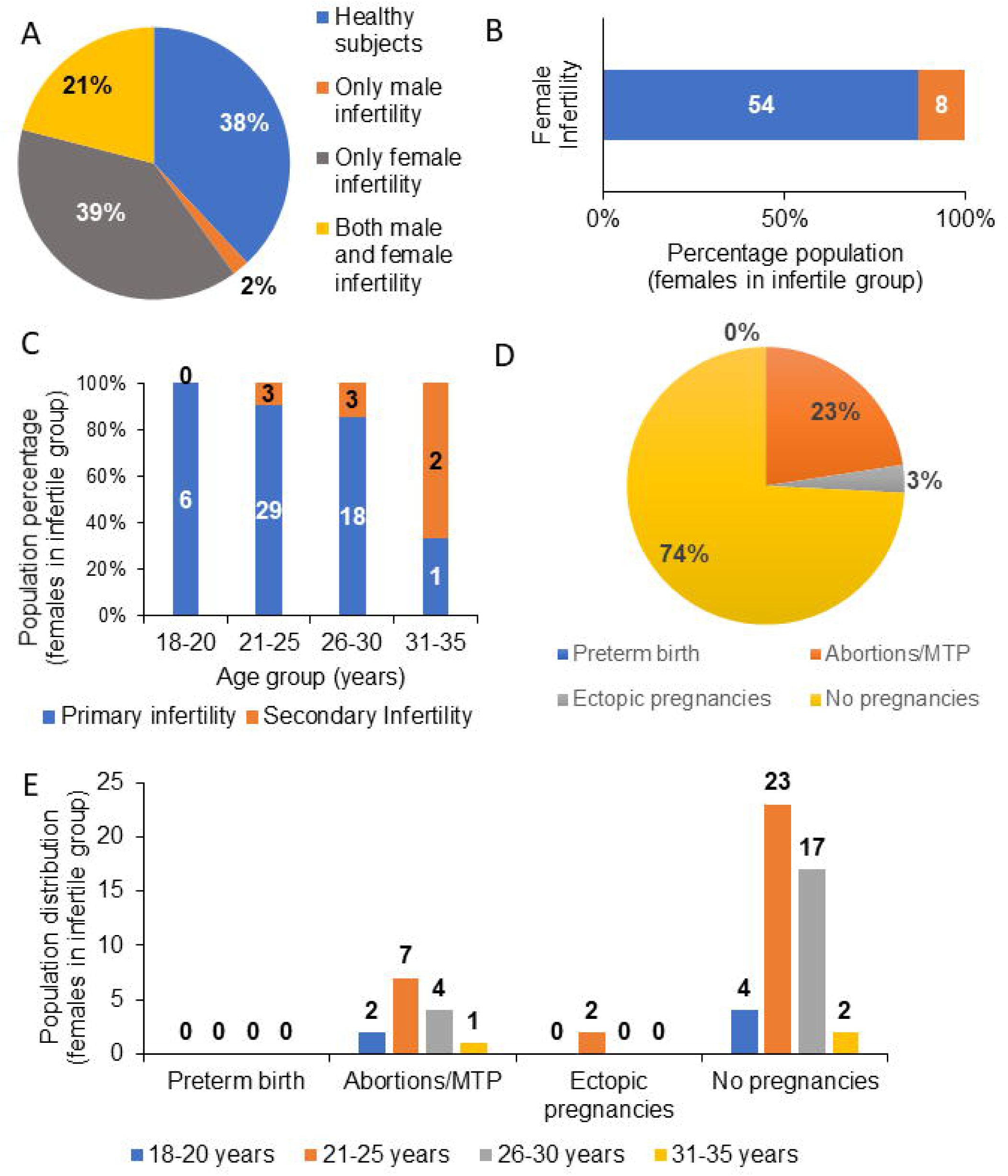
Population distribution and pattern analysis of female infertility and other reproductive conditions. A. Pie-chart shows the population distribution of infertility amongst the assessed subjects (n = 100, 62 infertile couples, and 38 healthy subjects). Out of 62 infertile couples, 60 cases were found to be associated with female infertility (39 cases of only female infertility and 21 cases of both male and female infertility), while 2 cases were identified as only male infertility leading to failure in conception. B. Histogram shows the percentage distribution of primary and secondary infertility amongst the 62 cases of infertility. C. Histogram shows the age-based percentage population distribution (females) of primary and secondary infertility amongst 62 cases of infertility. D. Pie chart shows the percentage population of preterm birth, spontaneous abortions/medical termination of pregnancy (MTP), ectopic pregnancy, and no conception amongst infertile females (n = 62). E. Age-based population distribution of females (n = 62) experiencing preterm birth, spontaneous abortions/MTP, ectopic pregnancy, and no conception.

### Genital infections-a major but neglected contributor to female infertility

Among the various parameters assessed in females belonging to the infertility group, incidence of genital infections (symptomatic and/or confirmed diagnosis) were thoroughly investigated in this study, such as vaginitis (vaginal itching, burning and discharge), PID, confirmed/symptomatic cases of UTIs (painful micturition/burning sensation on micturition), TB positivity (based on tuberculin test/ QuantiFERON-TB Gold/TB molecular diagnosis using PCR or TB culture test), cervicitis (cervical inflammation) and dyspareunia (painful intercourse) (**Table S1 and S2A**). Surprisingly, a large proportion of ∼27% of infertile females show one or more signs of infection-related symptoms (**Figure 3A, Table S2A**). The major symptoms observed in these subjects are vaginitis (∼18%), PID (∼26%), dyspareunia (∼11%), and 2 confirmed cases of pulmonary TB (∼3.2%) (**Figure 3B, Table S2A**). Interestingly, the analysis of questionnaire data reveals that confirmatory detection of infections has largely been overlooked and is not typically included in the routine check-ups involved in infertility management. The diagnosis is simply based on visible signs of genital infections listed above. Hence, the detection of genital infections may prove to be a highly valuable prognostic and diagnostic marker with significant relevance in the early management of female infertility.

**Figure 3.**
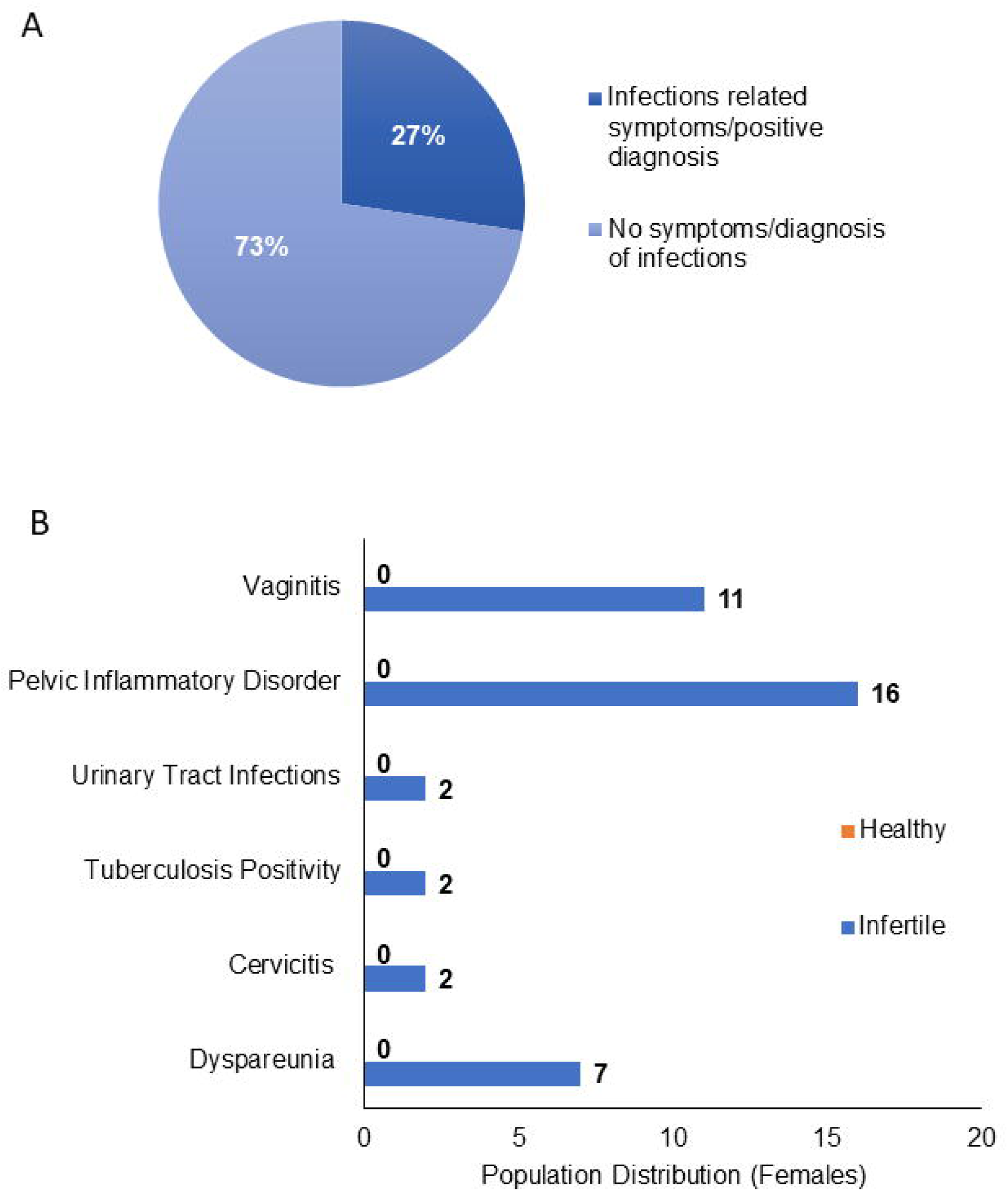
Contribution of genital infections to female infertility. A. The Pie chart shows the percentage of infertile females experiencing symptoms of genital infections or confirmed diagnosis of infection. Reports suggest tuberculosis infection (based on PPD tests) in 2 cases out of 62 subjects, while other infections are not identified during diagnosis. B. Histogram shows population distribution of various genital infection-related symptoms in females belonging to the infertile group (n =62, all cases of infertility) and healthy group (n = 38). Healthy females do not show any symptoms of infection, hence their bar is represented as “0”.

### Menstrual Health-a non-invasive prognostic marker of infertility

Menstrual irregularities appear to be the predominant symptoms observed in the case of females in the infertile group (∼42%, **Figure 4A, Table S2A and S2B**). Menstrual irregularities were further stratified into various categories such as menorrhagia (heavy and prolonged bleeding), dysmenorrhoea (painful periods with cramps), amenorrhoea (absence of periods), metrorrhagia (spot bleeding in between the periods) and oligomenorrhoea (irregular periods or inconsistent blood flow). Among these symptoms, amenorrhoea (∼13%), metrorrhagia (∼8%), and oligomenorrhoeae (∼15%) were evident only in the case of infertile subjects (**Figure 4B, Table S2A and S2B**). However, symptoms such as menorrhagia (∼34% vs. ∼24% in infertile vs. healthy) and dysmenorrhoea (∼65% vs. ∼29% in infertile vs. healthy) were observed both in healthy and infertile subjects, though a larger proportion is observed in the infertile group **(Figure 4B, Table S2A, and S2B)**.

**Figure 4.**
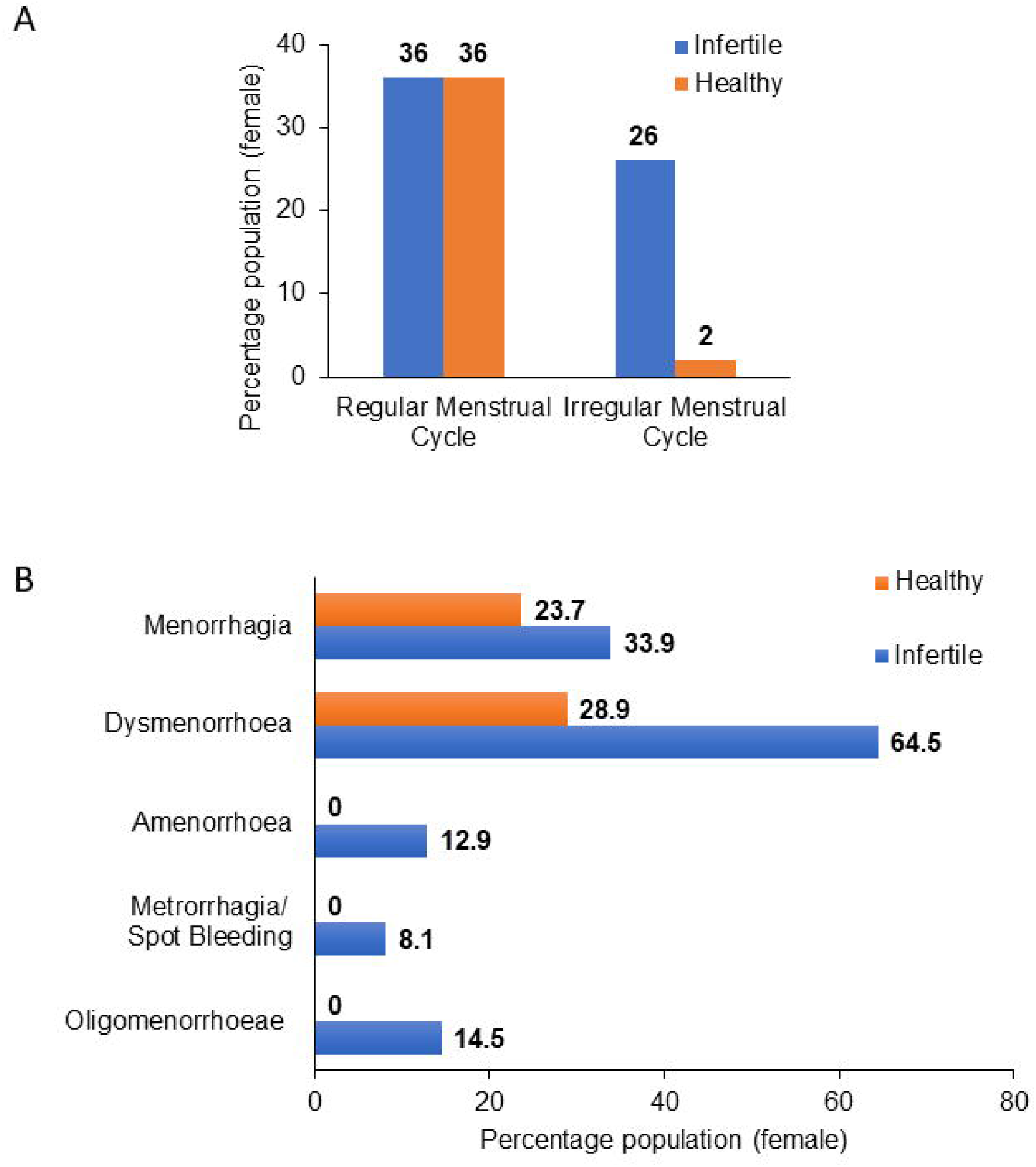
Female infertility and menstrual health. A. Histogram shows the percentage population distribution of females with respect to their menstrual health based on the regularity of the menstrual cycle. B. Histogram shows population distribution of various menstrual disorders among females belonging to infertile (n =62, all cases of infertility) and healthy groups (n = 38). Infertile [%] = [No. of patients with symptoms/Total infertility cases] *100. Healthy [%] = [No. of healthy subjects with symptoms/Total no. of healthy subjects] *100.

### Influence of physical health and lifestyle on infertility

Out of the 100 subjects, 63% of women report a lack of exercise in their daily routine. On further stratifying the data based on fertility status, a larger proportion of infertile females (n = 40, 65% of the infertile group) reported lack of exercise (**Figure 5A, Table S2A**). On further looking into the BMI status, all the females with low BMI (BMI < 18, 7% of the total study population) are infertile. Amongst the high BMI population, 10% are infertile (**Figure 5B, Table S2C**).

**Figure 5.**
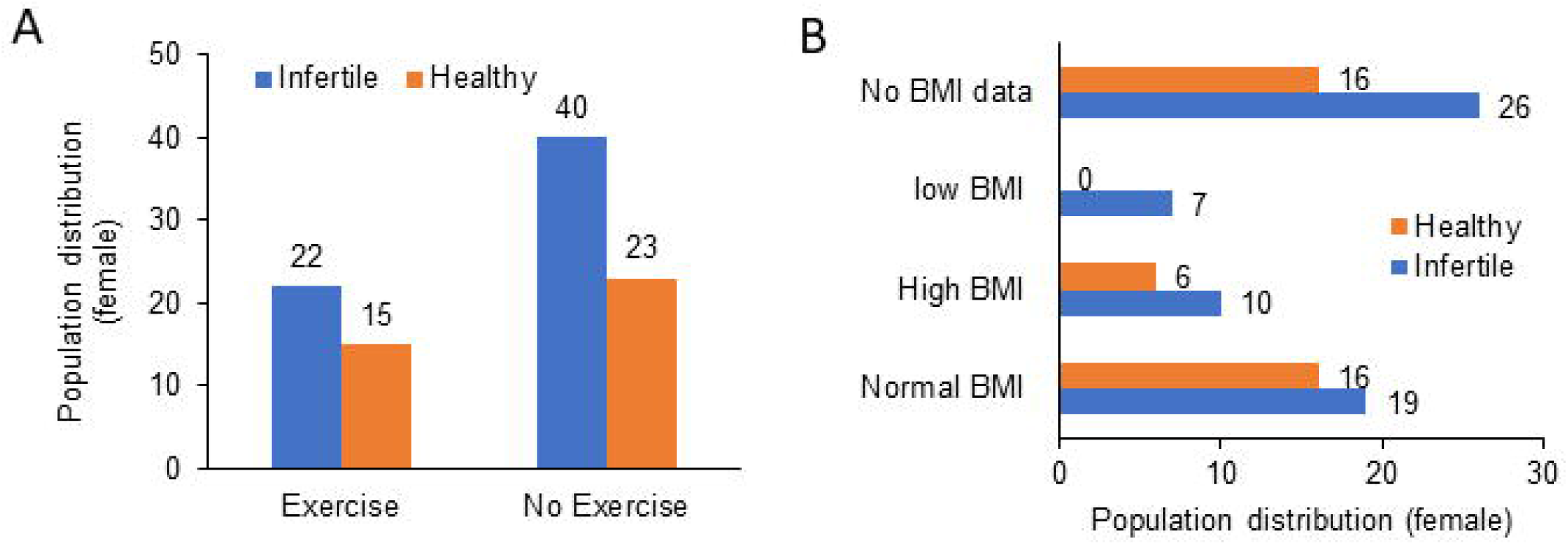
Contribution of lifestyle (exercise) and body mass index (BMI) to female infertility. A. Histogram shows the population distribution of all females (healthy and infertile) based on their exercise routine. B. Histogram depicts the population distribution of all females (healthy and infertile) with respect to their BMI. (Normal BMI:18-25; Low BMI: <18; High BMI > 25).

### Spectrum of reproductive tract anomalies in infertile females

We next assessed the spectrum of anomalies in the female reproductive tract in infertile subjects. Among the various anomalies, PCOS and PCOD are the most prominent pathological sequelae observed in infertile subjects (n = 19, ∼31%) (**Figure 6, Table S2A**). The other prominent symptoms include fallopian tube blockage (n = 5, ∼8%) and a variety of endometrial disorders, such as secretory/proliferative/thickened endometrium/hyperplasia/cysts (n = 4; ∼7%), and endometriosis (n = 2, ∼3%) (**Figure 6, Table S2A**). Unlike these symptoms, only one case of anovulation and ovarian teratoma is observed in the infertile group (**Figure 6, Table S2A**).

**Figure 6.**
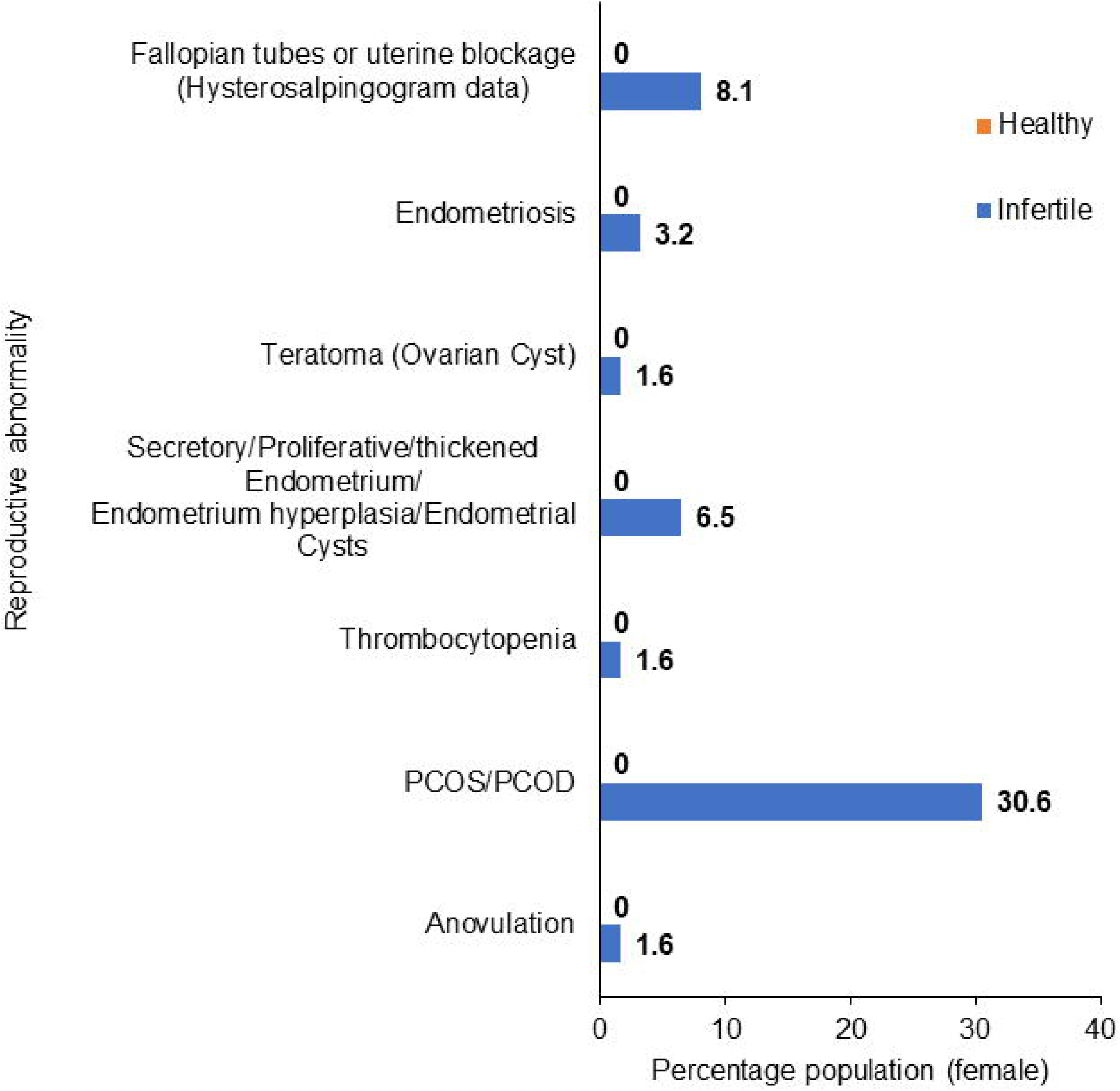
Female infertility and anatomical abnormalities of the reproductive tract. The histogram shows the population distribution of various anatomical abnormalities of the female reproductive tract in infertile (n =62, all cases of infertility) and healthy group (n = 38). Infertile [%] = [No. of patients with symptoms/Total infertility cases] *100. Healthy [%] = [No. of healthy subjects with symptoms/Total no. of healthy subjects]*100. Healthy females do not show any reproductive anomalies their bar is represented as “0”.

### Hormonal perturbations in infertile females

We next attempted to understand the changes in reproductive hormones in women experiencing infertility. We compared and categorized the hormonal profiles of the infertile subjects into low, high, and normal in the case of luteinizing hormone (LH) and follicle-stimulating hormone (FSH). In the case of anti-Mullerian hormone (AMH) and prolactin, data was stratified into abnormal (high) and normal. In the majority of the cases, data for hormone profiles is not available (**Table S2D**). Although 58% of the infertile subjects were tested for FSH, no specific pattern is observed concerning infertility (**Figure 7, Table S2D**). For other hormones, data is available only in < 23% of the infertile subjects, and no conclusive inference concerning fertility status could be drawn (**Figure 7, Table S2D**).

**Figure 7.**
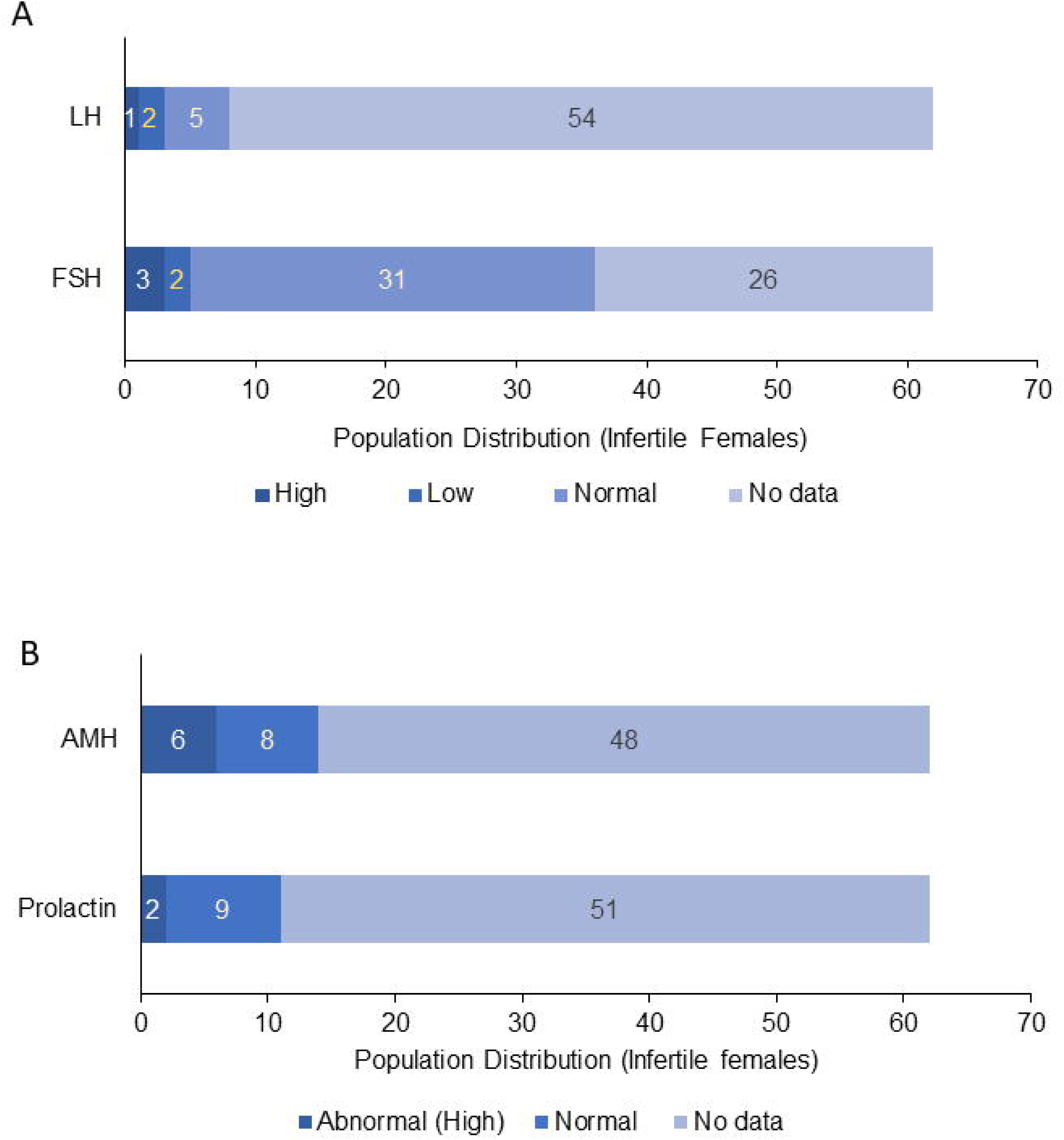
Female infertility and hormonal perturbations. The histogram shows the population distribution of females categorized under the infertile group (n = 62) experiencing abnormal (high/low) levels of female reproductive hormones such as A. FSH and LH; B. Prolactin and AMH. Hormone profiles of healthy subjects were not available and hence were not considered during analysis. FSH [Normal: 4-12 IU/L; high > 12 IU/L, low < 4 IU/L]; LH [Normal: 5-15 IU/L,; high > 15 IU/L, low < 5 IU/L]; Prolactin [Normal: ≤ 25 ng/ml; high > 25 ng/ml]; AMH [Normal: < 3 ng/ml; High > 3 ng/ml]

### Male infertility and semen abnormalities in the infertile group

Among the 62 infertile couples, a large proportion of male counterparts (n = 23, 37%) presented with a variety of semen abnormalities (**Figure 8A**), such as, oligospermia (low sperm count), low sperm DNA integrity (SDI), asthenozoospermia (AS) or low sperm motility, teratozoospermia (high TZI index > 1.6) indicating presence of abnormal shaped sperms (often linked with infection, injury or smoking), pyospermia (presence of pus cells in semen), hypospermia (low volume of semen) and hyperviscosity (**Figure 8B, Table S2A**). The most predominant abnormalities are oligospermia (21%), asthenozoospermia (23%), and teratozoospermia (∼ 15%). A few cases of pyospermia (8.1%) and hyperviscosity (5%) are also observed indicative of infection or high levels of seminal leukocytes (**Figure 8B, Table S2A**) also indicated previously [26, 27].

**Figure 8.**
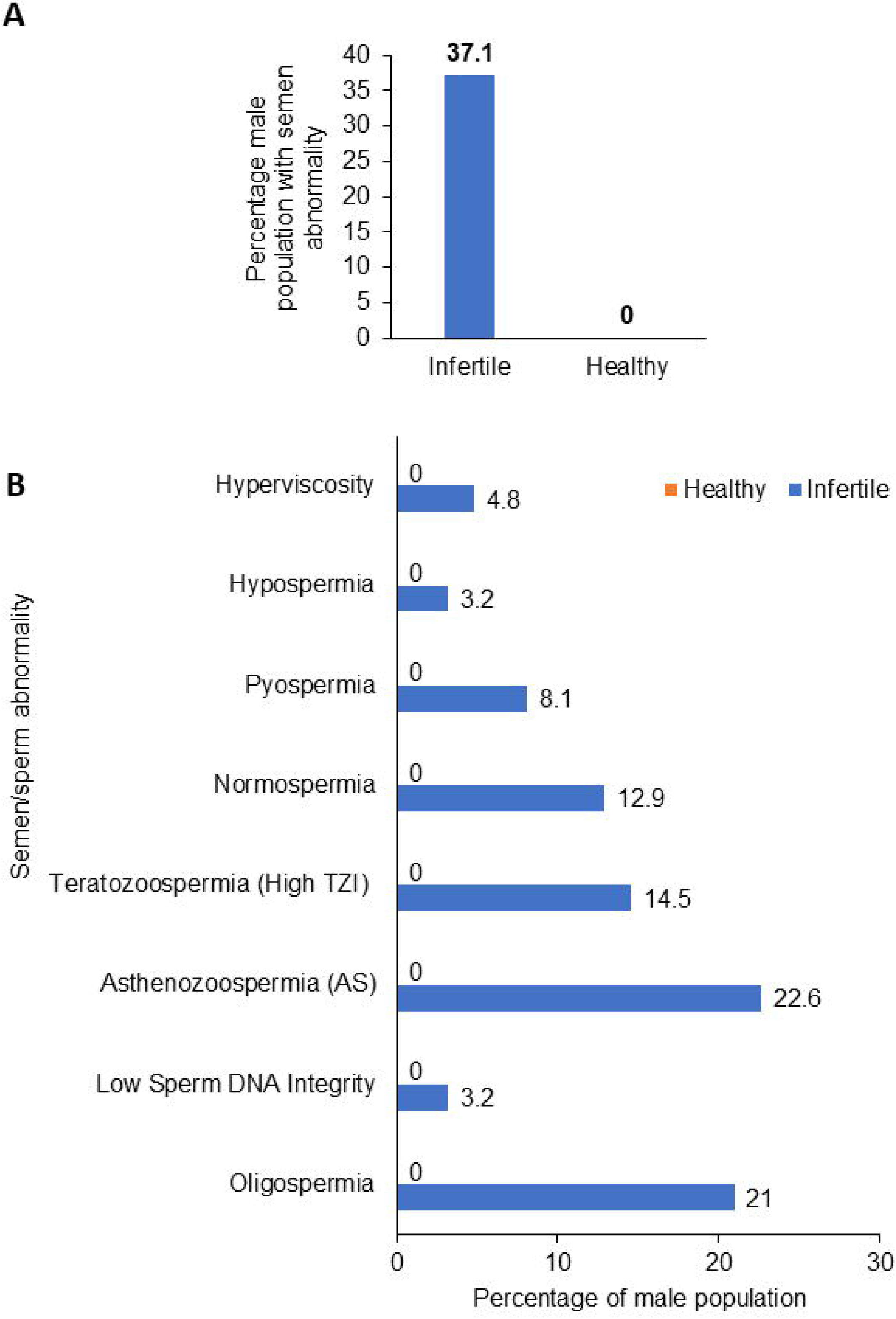
Male infertility and semen abnormalities. A. The histogram shows that male infertility is primarily diagnosed with abnormal semen analysis in the infertility group (n = 62) and healthy group (n = 38). B. Histogram shows the various semen abnormalities prevalent in male infertile patients in the study. Infertile [%] = [No. of male subjects with symptoms/Total infertility cases]*100. Healthy [%] = [No. of healthy subjects with symptoms/Total no. of healthy subjects]*100. Males of healthy group do not show any semen abnormalities and hence in absence of data, their bar is represented as “0”.

### Multivariate correlation analysis among various parameters

The major objective of this study is to identify the most significant parameters (**Table S1**) that impact fertility and reproductive health in females. We also wanted to assess if changes in these critical female parameters co-occur/correlate with the reduced sperm/semen quality in males. We stratified the parameters into various categories such as (a) menstrual disorders (**Table S2A and S2B**), (b) reproductive tract anomalies such as endometriosis, other endometrial disorders (e.g. thickened/secretory/proliferative endometrium/hyperplasia/cysts), fallopian tube or uterine blockages, PCOS/PCOD, anovulation and ovarian cysts/teratoma (**Table S2A**), (c) genital infection associated symptoms/confirmed diagnosis of infections like TB/UTIs (**Table S2A**), (d) physiological parameters such as reproductive hormones (FSH, LH, Prolactin, AMH) (**Table S2D**), (e) physical parameters and lifestyles, such as weight, height, BMI and exercise routine (**Table S2A and S2C**) and (f) male sperm and semen abnormalities (**Table S2A**). To derive the correlations between various parameters, we performed multivariate correlation analysis (Phi correlation) [28–30].

We observe a highly significant and very strong correlation between parameters related to genital infection-associated symptoms (or confirmed diagnosis) and menstrual disorders such as oligomenorrhoea, amenorrhoea, metrorrhagia, and dysmenorrhoea. (**Figures 9 and 10**). Further, a very strong and significant correlation was observed between genital infections and perturbations in the female reproductive hormones such as LH **(Figures 9 and 10)**. The presence of infection-associated symptoms also is strongly correlated with the occurrence of PCOS and PCOD, uterine and tubal blockage, ovarian cysts, and endometrial disorders **(Figures 9 and 10)**.

**Figure 9.**
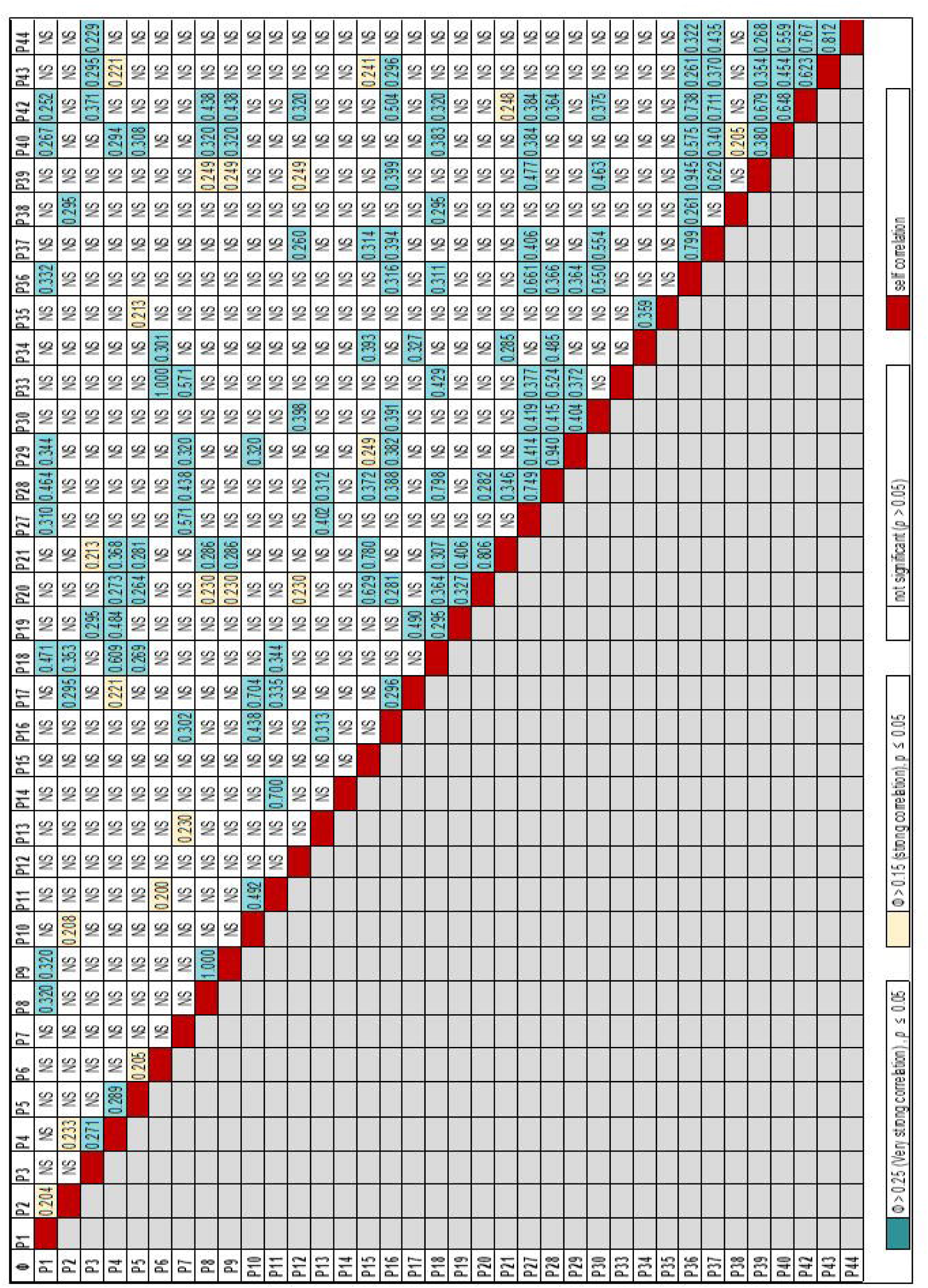
Correlation heat map between multiple variables. The data gathered by a questionnaire-based assessment of female patients is used for finding out the correlation amongst the various parameters used for the assessment. A multivariate correlation value (Phi correlation with the significance of correlation *p* ≤ 0.05) was used for generating this correlation heat map. (P1-P45 are the various parameters assessed in this study, for description see supplementary table S1).

**Figure 10.**
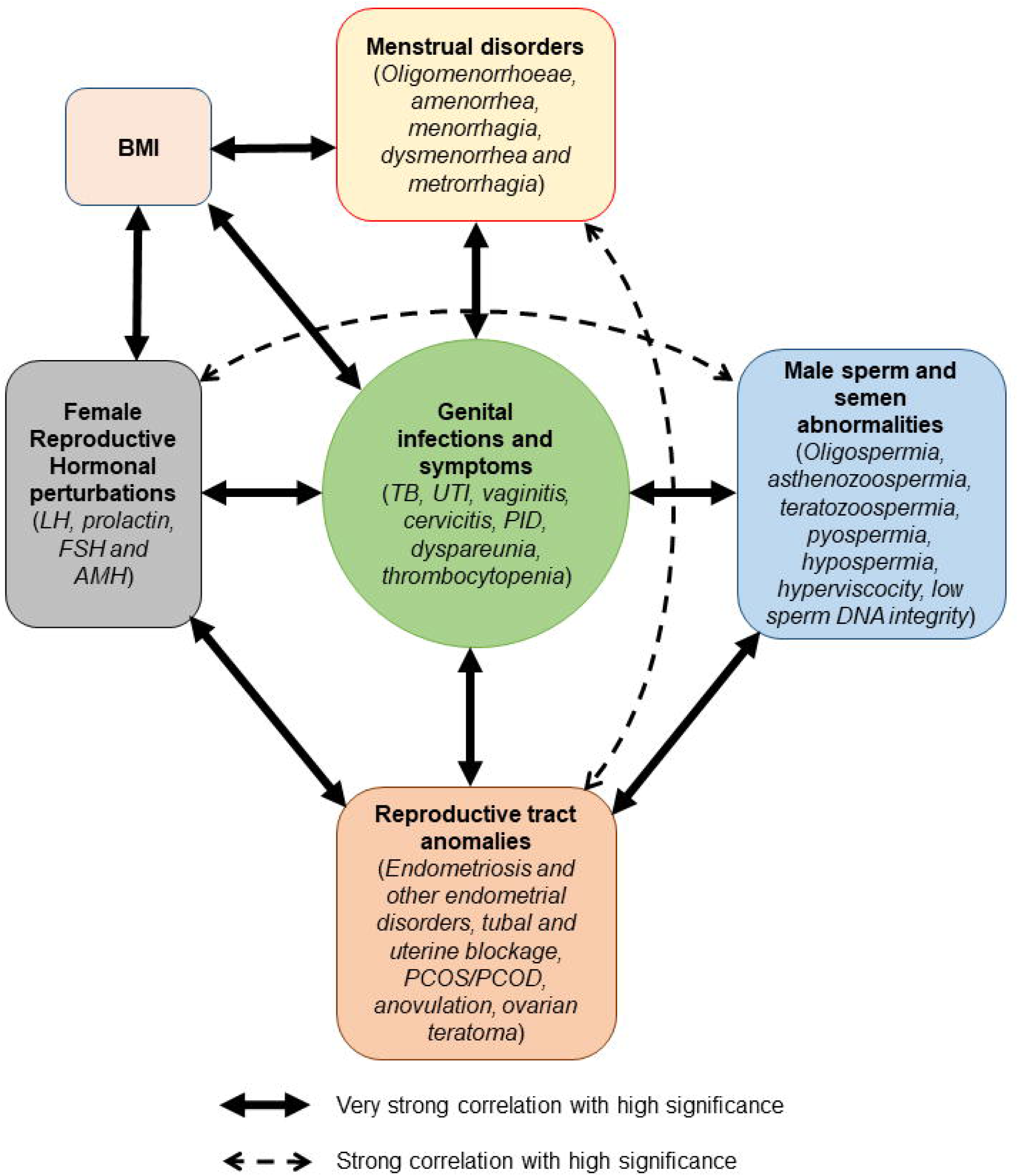
Schematic diagram showing correlation between various fertility parameters and their association with the incidence of genital infections. The correlation between various parameters is depicted schematically with the help of bidirectional arrows of varying thickness corresponding to the strength of correlation between these parameters. The correlations are deduced from the multivariate correlation analysis (Phi correlation and the significance of the correlation). For details, refer to the correlation heat map shown in Figure 9.

We next assessed if female genital infections correlate/co-occur with abnormalities in the male semen and sperm profile. Our study suggests a very strong and significant correlation of female genital infections with the occurrence of teratozoospermia, pyospermia, and hyperviscosity of semen and a strong correlation with asthenozoospermia. The abnormalities observed in male partners of infertile females in this study concur well with the previously reported sperm abnormalities during male genital infections [26, 27], however, no confirmatory diagnostic reports were available to verify the nature of infections in these males (**Figure 9 and 10**). Another key finding is a very strong and significant correlation between male semen/sperm abnormalities characteristic of infections, such as low sperm DNA integrity, asthenozoospermia, teratozoospermia, pyospermia and hyperviscosity with female disorders such as occurrence of PCOS and PCOD, uterine and tubal blockage, oligomenorrhoeae, amenorrhoea, metrorrhagia & dysmenorrhoea (**Figure 9 and 10**). These observations highlight that transmission of genital infections between sexual partners could potentially influence female menstrual health and fertility in both males and females. Male factors are also strongly correlated with hormonal perturbations in infertile females such as FSH, LH, and AMH (**Figures 9 and 10**).

Among other parameters, we observe a strong and significant correlation between reproductive tract anomalies such as PCOS and PCOD with the occurrence of menstrual disorders such as amenorrhoea (**Figures 9 and 10**). We also observe endometrial disorders to impact menstrual health and are significantly correlated with menorrhagia. We next assessed if the reproductive tract anomalies are linked with reproductive hormones. We observe a strong and significant correlation between ovarian cysts with AMH and anovulation with FSH, LH, and prolactin (**Figures 9 and 10**). In addition, uterine and tubal blockage are strongly correlated with prolactin, LH, and AMH levels (**Figures 9 and 10**). However, one limitation of the data collected was the absence of reports corresponding to female reproductive hormones in a large number of cases (**Figure 7, Table S2D**).

Peculiarly, we observe a very strong and significant correlation between BMI in females and genital infection-associated symptoms (such as dyspareunia/cervicitis/vaginitis), menstrual disorders (such as menorrhagia), and reproductive hormones (such as LH) (**Figure 9 and 10**).

## Discussion & Conclusion

Remarkably, our investigation revealed that genital infections are often overlooked during pregnancy or in women of reproductive age, indicating a glaring gap in current healthcare practices and a significant lack of robust point-of-care (POC) diagnostic tools in rural areas for detecting genital infections, including TB. Hence, infertile women are typically treated primarily with hormones (Letrozole or clomiphene) to improve ovulation, hormonal stimulation, and improvement of menstrual health and are not treated for infections unless there are visible signs. As a result, asymptomatic females remain undiagnosed for infections and do not receive appropriate health care promptly. This prolonged lack of diagnosis and treatment can lead to chronic infections and consequential pathological sequelae, ultimately resulting in infertility.

One of the most striking revelations from our study was the early onset of infertility, occurring between 18 to 25 years of age. This suggests that many affected individuals might have experienced mild or chronic symptoms post-puberty, which remained undiagnosed and untreated. Importantly, a substantial portion of these patients exhibited classical signs of infection (27%), while 73% were rendered ineligible for confirmatory diagnostic tests due to a lack of classical symptoms of infections. This underscores the urgent need for more comprehensive screening strategies. We then uncovered a range of reproductive tract anomalies in infertile females, including signs of PCOD/PCOS, tubal blockage, and smaller percentages with conditions such as endometriosis, anovulation, uterine fibroids, and ovarian cysts. This diversity of conditions makes it challenging for clinicians to establish a clear link between these disorders and genital infections.

Furthermore, we observed a very strong and significant correlation between genital infections and both physiological (hormonal profiles and BMI) and pathological effects on female reproductive health (reproductive anomalies and menstrual disorders) and sperm/semen quality in the case of males. Hence, detecting and treating these infections is crucial not only for improving female reproductive health but also for enhancing the success rate of fertility treatments, including assisted reproductive technologies (ART). This suggests a potential link between poor lifestyle habits and these health issues, similar to associations found with diabetes, obesity, and PCOS/PCOD [31, 32].

Diagnosing genital infection in case of TB, BV, candidiasis, PID, and endometritis can be challenging due to varying, mild, or absent symptoms [13]. Moreover, identifying upper reproductive tract pathogens (endometrial/tubal/ovarian) through microbial culture could be challenging requiring invasive sampling and there is also the risk of vaginal and/or cervical pathogens contaminating these samples [13, 20–22]. Therefore, a modest clinical suspicion of genital infection is critical to identify women at risk to prompt them for further detailed clinical investigations [33]. The comprehensive questionnaire or survey-based approach as designed by us in this study could address this gap and help in patient stratification for downstream confirmatory diagnosis and treatments. On confirmatory diagnosis, treatment for infections using suitable antimicrobials has shown benefits in terms of improving symptoms and fertility outcomes [20–22, 34, 35]. Asymptomatic patients with chronic endometrial disorders and unexplained infertility, recurrent implantation failures, miscarriages, or intrauterine infections in previous pregnancies (like chorioamnionitis or deciduitis), should also be considered for confirmatory diagnosis for infections.

Hence, we stress the potential of utilizing questionnaires for self/clinical assessment, possibly through survey forms/user-friendly phone or web applications in regional languages. This innovative strategy will also empower women from various backgrounds to report reproductive health issues at an early stage, leading to timely diagnosis and treatment thereby preventing chronic infections and long-term damage in the form of infertility. This may also prevent medical poverty associated with exorbitant treatment costs linked with ART. Our study also highlights an urgent need for improvement in point-of-care (POC) diagnostic tools, increased awareness, and a more holistic approach to tackle genital infections in the context of infertility. By bridging these gaps, we can make significant strides in enhancing reproductive health outcomes for women in rural and semi-urban settings, especially low and middle income countries, promoting overall well-being and fertility.

## Author Contribution

NP and RJD have conceived and designed the project, performed data mining, analyzed the results, designed figures as well written and edited the manuscript. NP collected the data at the clinic. US (epidemiologist) performed the statistical analysis. RJD (microbiologist and biochemist), MJ (medical microbiologist), and VH (OB & Gynaecologist, Curewell Hospital) provided overall supervision and coordination during data collection at the clinical site. VH contributed to the enrolment of subjects and patient stratification based on their fertility status. RJD received the funding. All the authors reviewed and edited the manuscript.

## Acknowledgments

RJD and NP are thankful to Amancha Sireesha for technical support and to the medical staff at Curewell Hospital for support during the primary assessment and stratification of subjects.

## Data availability statement

All data that supports the findings of this study are available from the corresponding author upon reasonable request.

## Supplementary Materials

Supplementary materials contain the following:

**Supplementary Material 1 – Patient consent form**

**Supplementary Material 2 – Patient Questionnaire**

**Supplementary Material 3 - Tables S1-S2 (A, B, C and D)**

