## Supplementary Materials for "A cross-sectional questionnaire-based landscaping of female infertility reveals genital Infections as a major contributor to reproductive tract anomalies, menstrual disorders, and infertility"

**List of Supplementary Materials**

- **Supplementary Material 1 – Patient consent form**
- **Supplementary Material 2 – Patient Questionnaire**
- **Supplementary Material 3 - Tables S1-S2 (A, B, C and D)**

### Supplementary Material 1 – Patient consent form

#### 1. Patient Declaration/Informed Consent Form (English)

**I declare that the information provided by me is true and correct to the best of my knowledge and belief. I provide my consent to be contacted for further clarifications/discussions.**

I have been asked to take part in a research study *"Design and development of rapid and cost-effective point-of-care, nano-based nucleic acid biosensors for prognosis, diagnosis and management of infection-induced infertility in females."* carried out by **BITS-Pilani, Hyderabad campus** in collaboration with **Curewell Hospital, Warangal**. The study team has described the following details pertaining to the research topic with special emphasis on its benefits for the maternal and child health. (The details have been explained to me in English/Telugu/Hindi).

a. **Why the study is being done?** [The study will be helpful to find out the prevalence of genital infections and its association with reproductive tract disorders including infertility in females in the region of Warangal, Telangana]
b. **What will happen to me if I participate in the study?** [Clinician will be able to diagnose the infections on time and will be able to decide the most appropriate treatment specific to a particular infection(s) which will provide effective and timely symptomatic relief and prevent worsening of the symptoms.]
c. **How long I will be in the study?** [It is going to be a one-time interaction on enrolment, where you will fill a questionnaire and provide consent to give urine/vaginal swab samples as part of routine procedure in the hospital for regular diagnosis. The leftover samples (waste) will be given to us for our study]
d. **The possible risks, discomforts, and the benefits of the study?** [Since the study involves minimally invasive techniques for self-sampling/sample collection by a trained medical technician at the hospital, there is minimal to no risk or discomfort involved. Plus, the study will be beneficial for research related to development of cost effective diagnosis methods important for maternal and fetal health]
e. **How my study records will be kept private?** [Multiple steps will be taken to ensure data anonymization/de-identification/security, such as
   1. The identities of the patients will not be disclosed in any publication/conferences/reports to funding agency/public forum or to any commercial organization.
   2. Patients samples will be double blinded and assigned codes – one at the level of clinic for the clinician and will also be de-identified and coded for research purpose. These codes will be alphanumeric in nature and assigned at hospital by the investigating clinician and will be then de-identified and coded when sent to PI's lab for research purpose.
f. **Whether the study will cost me anything?** [The patient will not be charged anything for this study]

I may contact Doctors of the research team at any time if I have questions about the research. In future I have the liberty to withdraw from the study at any time. I, hereby, give my consent to provide my details in the questionnaire and subsequently, give my consent to use the leftover urine/vaginal swab samples which are collected by the clinician as part of routine diagnosis to be used for the aforementioned research study.

| Patient Name | Patient's Signature* | Date |
|---|---|---|
| ________________________ | | ____________ |
| Reviewed by | Reviewer signature | Date |
| _________________________ | | ____________ |

### Supplementary Material 2 - Patient Questionnaire

| For office use only |
|---|
| **Patient Details:**<br><br>**Name:**<br>**Gender:**<br>**Age:**<br>**Contact no.:**<br>**Address: ______________________**<br><br>**Patient ID (Hospital):** |
| **Patient ID (De-identified): (generated slip)** |

---

#### Questionnaire for the female patient

#### I. Personal Information

| | |
|---|---|
| 1. | Partner's Age |
| 2. | Patient Occupation |
| 3. | Partner's Occupation |
| 4. | Reason for the visit to clinic |

#### II. General Health Details (Female patient)

| | |
|---|---|
| Height (cms) = | Weight (kgs) = |
| Blood type = | |
| Do you exercise/workout?<br>Yes - No - | List of exercises and duration |
| Any addictions (caffeine/tobacco/alcohol/drug abuse):<br>Yes - No - | |

#### III. Menstrual History

| | |
|---|---|
| 1. Age at first period? | ___________yrs |
| 2. First day of your last period? ___________ | Normal_______ / Abnormal__________ |
| 4. Are your periods regular?<br>Yes ______ /No __________ | Mention the gap between the periods: |
| 5. Details of menstrual flow. | Light_____ /Moderate_____ /Heavy_____ |
| 6. Duration of the flow | _____________days |
| 8. Do you experience cramps before, during, or after your period? Yes ____ No _____ | If yes: Cramps are<br>Mild______ Moderate_____ Severe______<br><br>Medication for cramps (if taken)<br>_____________________________________ |
| 9. Do you bleed or spot between periods? | Yes ______/ No_________ |

IV. **Pregnancy History**

**a) Did you have term pregnancies previously? Yes - No-**
**If yes, then how many**
**b) Did you have preterm pregnancies/deliveries previously? Yes- No-**
**If yes, then give details of health of the delivered baby**
**c) Did you have natural abortions earlier? Yes- No-**
**If yes, then how many ___________**
**d) Did you have medical termination of pregnancy (medical abortions) previously? Yes- No-**
**If yes, then how many ___________**
**e) Did you have Ectopic pregnancy/tubal pregnancy previously? Yes- No-**
**If yes, then how many ___________**

V. **Contraceptive / Sexual History**

| | |
|---|---|
| 1. What form of contraception do you use now or have you used in the past? (Tick wherever necessary.) | Pills / IUD / Diaphragm Rhythm<br><br>Withdrawal Foams/Jellies/Condom<br><br>Tubal ligation / Vasectomy |
| **2.** If you've ever been on oral contraceptives(pills), were your periods regular after stopping the pills?<br>**Yes_______ No___________** | |
| **3.** Do you time intercourse around ovulation?<br>**Yes________ No ________**<br>_______________________________________________ | If yes, how? |

VI. **Fertility Treatment History**

1. Have you been treated for infertility before? **Yes_____ No________**
If yes, who was your physician? ______________________________________________________________
Diagnosed cause? ___________________________________________________________________________

| **Treatment History** | | | |
|---|---|---|---|
| Intrauterine insemination | # Cycles _ | Dates / to __/ | Outcome: Pregnant/ Delivered/ Ectopic/ Miscarriage/ Not pregnant |
| Clomid/Letrozole alone maximum tablets/day? _ | | / to __/ | Outcome: Pregnant/ Delivered/ Ectopic/ Miscarriage/ Not pregnant |
| Clomid/Letrozole with intrauterine insemination | | / to __/ | Outcome: Pregnant/ Delivered/ Ectopic/ Miscarriage/ Not pregnant |
| Daily hCG injections/ intrauterine insemination maximum # vials/day? _ | | / to __/ | Outcome: Pregnant/ Delivered/ Ectopic/ Miscarriage/ Not pregnant |
| Completed IVF cycles | | / to __/ | Outcome: Pregnant/ Delivered/ Ectopic/ Miscarriage/ Not pregnant |
| Frozen embryo cycles | | / to __/ | Outcome: Pregnant/ Delivered/ Ectopic/ Miscarriage/ Not pregnant |
| Canceled IVF attempts | | / to __/ | |
| Any other treatment? (Describe) | | | |
| **Current Diagnosis (Tick whichever is applicable):** | | | |

**1. Amenorrhoea/Dysmenorrhoea/Irregular periods/Spot bleeding**

**2. Abortions/MTP**

**3. PCOD(PCOS)/ PID (Pelvic Inflammatory Disease)/ Endometriosis**

**4. Hyperthyroidism / Hypothyroidism**

**5. Do you experience any of the following symptoms (Tick whichever is applicable):**
**a) Vaginal Itching/burning. (Yes- No- )**
**b) Inflammation or soreness of vagina (Yes- No- )**
**c) Vaginal Discharge (Yes- No- )**
**d) Painful micturition (pain during urination): (Yes- No- )**
**e) Pain during vaginal sex: (Yes- No- )**
**f) Vaginal bleeding or spotting during vaginal sex: (Yes- No- )**

2. Any previous history of following ailments? (Tick whichever is applicable)

| | | | |
|---|---|---|---|
| High Blood pressure/ Low Blood pressure | Diabetes | Epilepsy | Vertigo |
| Heart diseases | Neurological Conditions | Kidney diseases | Liver diseases |
| Hyperthyroidism/ Hypothyroidism | Chronic Bronchitis | Chronic Migraines and headaches | Bowel irritation |
| Urinary tract infection | Gonorrhoea | Chlamydia | Syphilis |
| Hepatitis | Tuberculosis | Measles | Pneumonia |
| Sexually Transmitted Diseases (HIV, HPV, HCV) | Vaginal Candidiasis | Trichomoniasis | Cancers/Tumors |

Any allergies (Food allergen/Environment allergen/Drug/ Antibiotic allergens/autoimmune disorders)
Yes- /No- _______Mention the name________________________________________________________

3. What drugs are you taking as part of current treatment procedure?

| | |
|---|---|
| Letrozole (Femara) | Clomiphene citrate (Clomid) |
| hCG (Profasi, Pregnyl, Novarel) | HMG (Repronex, Pergonal, Menopur) |
| bromocriptine (Parlodel) | Progesterone |
| Estrogen | Danazol/Danocrine |
| Prednisone | FSH (Gonal F, Follistim) |
| GnRH agonists (Lupron, Synarel) | Others? Specify. |

4. Which of the following tests have you had performed? Check all that apply and the results if known:

| | Values | Results? |
|---|---|---|
| Pap smear | | |
| Basal Body Temperature (BBT) charts | | |
| **Hormonal Tests** | | |
| Follicle stimulating hormone (FSH) | | High /Low /Normal |
| Leutenising hormone (LH) | | High /Low /Normal |
| Prolactin | | High /Low /Normal |
| Anti-Mullerian Hormone (AMH) | | High /Low /Normal |
| Estrogen | | High /Low /Normal |
| Progesterone | | High /Low /Normal |
| DHEA-S | | High /Low /Normal |
| Testosterone | | High /Low /Normal |
| Thyroid tests | Yes /No | High /Low /Normal<br><br>Are you on medication; If yes (name and dosage) |
| **Radiological/Invasive Examination** | | |
| Endometrial Biopsy | Yes /No | |
| Hysterosalpingogram (X-ray of uterus/tubes) | Yes /No | |
| Ultrasound/Sonogram | Yes /No | |
| Laparoscopy | Yes /No | |
| **Examination of genital infections** | | |
| Chlamydia/Gonorrhoea test | Yes /No | |
| Urinary tract infection. | Yes /No | If yes, provide the name of infection: |
| PPD/Quantiferon TB test/TB PCR test/TB culture test? | Yes /No | Results:<br><br>If yes, are you on TB treatment/DOTS - _________ |
| Others infections? Specify. | | |

5. **Past surgery(s)/treatment(s)**

| **Type of surgery** | **When?** |
|---|---|
| Surgery for reversal of tubal ligation? **Yes / No** | |
| Surgery for pelvic adhesions or other pelvic surgery?<br>**Yes / No** | |
| Treatment of cervix for abnormal Pap Smear? **Yes / No** | |
| Other surgeries (D&C, ovarian, appendectomy, thyroid)?<br>**Yes / No** | **Specify details:** |
| If you've had anaesthesia, have you had problems with anaesthesia?<br>**Yes / No** | **Specify details:** |

### VII. Partner's health history:

| | |
|---|---|
| Has your partner had a semen analysis?<br>**Yes /No** | **Normal /Abnormal** |
| **Sperm Analysis stats** | |
| Sperm count:<br>Sperm motility:<br>TZI (Teratozoospermia index) levels: | UTI test:<br>White blood cells: |
| Is your partner seeing a doctor for evaluation of infertility? **Yes / No** | If yes, what is the diagnosis and how is he being treated? |
| Has he ever fathered a child previously, either with you or with other women?<br>**Yes /No** | If yes, when? |
| Is your partner ever diagnosed with Tuberculosis/UTI/STD? **Yes /No** | If Yes, mention details and prescribed drugs/duration of therapy |

### VIII. Family History

| | |
|---|---|
| **1.** Did your mother have any difficulty with conception or pregnancy or abortions or pre-term birth?<br>**Yes____ No____** | **If yes, give details.** |
| **2.** Is there a family history of infertility?<br>**Yes____ No _____** | If yes, who (list all members and relationship to you): |
| **3.** Is there a family history (mother and father side of the female patient)?<br>Bleeding tendencies / Strokes / Cancer / Thyroid Disorders / Diabetes / Tuberculosis /<br>(HPV, HCV, HIV) / Heart Disease / High Blood Pressure<br>Other (Specify) ____________________________________________ | |

### Supplementary Material 3 – Tables

**Table S1. List of parameters.** Table enlists the details of parameters used for the questionnaire based assessment of the subjects. Each parameter is provided with a variable name (ID), starting from P1 to P45. The IDs of the variable/parameter are assigned for the ease of statistical analysis – frequency distribution and Phi correlation.

| ID | Parameter |
|---|---|
| P1 | Oligomenorrhoeae / Irregular Periods or inconsistent blood flow |
| P2 | Polycystic ovarian syndrome (PCOS)/Polycystic ovarian disease (PCOD) |
| P3 | Metrorrhagia/Spot Bleeding |
| P4 | Amenorrhoea/Absence of periods |
| P5 | Dysmenorrhoea/Painful periods with cramps |
| P6 | Menorrhagia (Heavy Bleeding) |
| P7 | Anovulation |
| P8 | Vaginal discharge/leukorrhea |
| P9 | Thrombocytopenia |
| P10 | Tubal Blockage |
| P11 | Secretory/Proliferative/thickened Endometrium/Endometrium hyperplasia/Endometrial Cysts |
| P12 | Teratoma (Ovarian Cyst) |
| P13 | Abortions/Medical termination of pregnancy |
| P14 | Endometriosis |
| P15 | Dyspareunia (pain during intercourse) |
| P16 | Fallopian tubes or uterine blockage |
| P17 | Cervicitis (Cervical inflammation) |
| P18 | TB Positivity (PPD/Quantiferon TB test/TB PCR test/TB culture test) |
| P19 | Confirmed/Symptoms of UTI (painful micturition) |
| P20 | Pelvic Inflammatory Disorder |
| P21 | Vaginitis/symptoms of infections (Itching, burning, discharge) |
| P22 | Treatment with Clomid and/or Letrozole alone |
| P23 | Treatment with Estrogen Priming followed by Letrozole and/or Clomid |
| P24 | Treatment with Letrozole/Clomid priming followed by Progesterone |
| P25 | Treatment with Estrogen Alone |
| P26 | Treatment with Progesterone Alone |
| P27 | Follicle stimulating hormone (FSH) levels |
| P28 | Luteinising hormone (LH) levels |
| P29 | Prolactin hormone levels |
| P30 | Anti-Mullerian Hormone (AMH) levels |
| P31 | Age at first period? (in years) |
| P32 | Duration of the menstrual cycle (in days) |
| P33 | Details of menstrual flow? |
| P34 | Body Mass Index (BMI) Status |
| P35 | Do you exercise or workout? |
| P36 | Was the semen abnormal? |
| P37 | Oligospermia (Low Sperm Count) |
| P38 | Low Sperm DNA Integrity |
| P39 | Asthenozoospermia (AS) or Low motility Sperm |
| P40 | High TZI (Teratozoospermia index) levels |
| P41 | Normospermia |
| P42 | Pyospermia [Presence of pus cells] |
| P43 | Hypospermia [Low volume of semen] |
| P44 | Hyperviscosity |
| P45 | Treated for TB with Macoz (Rifampicin and Isoniazid) |

**Table S2. Frequency distribution analysis of parameters in infertile and healthy groups.** The table represents frequency distribution of individuals in each population group (healthy and infertile) with respect to parameters. A. Pathological sequelae- menstrual disorders (P1, P3-6) and reproductive tract anomalies (P2, P7, P10-14 and P16), genital infection associated symptoms or confirmed diagnosis of infections (P8, P9, P15, P17-21), female treatment (P22-26), exercise (P35), male reproductive semen/sperm abnormalities (P36-44) and male treatment (P45); B. Details of menstrual flow (P33) and C. Body Mass Index (BMI - P34). D. Female reproductive hormones (P27-30)- FSH, LH, Prolactin and AMH. The percentage frequencies for each parameter (or variable) are calculated with respect to the total frequencies in the respective group (infertility group: 62 and healthy control group: 38). Frequency distribution in all these parameters are analysed on the basis of multinomial response "Yes" (represented as 1) and "No" (represented as 0) and absence of response by the subjects for any parameter is recorded as "No data". P31 and P32 are excluded from the frequency analysis as the responses for these parameters are continuous in nature.

**A.**

| **Category of parameter** | **ID** | **Parameter** | **Total frequency of infertility group, N1 = 62** | | **Total frequency of healthy control group, N2 = 38** | | ***Remarks about the parameters (if any)*** |
|---|---|---|---|---|---|---|---|
| | | | **Frequency of patients (F1)** | **Infertile [%] (F1*100/N1)** | **Frequency of healthy subjects (F2)** | **Healthy [%] (F2*100/N2)** | |
| **Pathological sequelae (Menstrual disorders)** | P1 | **Oligomenorrhoeae / Irregular Periods or inconsistent blood flow** | 9 | 14.5 | 0 | 0 | |
| | P3 | **Metrorrhagia/ Spot Bleeding** | 5 | 8.1 | 0 | 0 | |
| | P4 | **Amenorrhoea/Absence of periods** | 8 | 12.9 | 0 | 0 | |
| | P5 | **Dysmenorrhoea/ Painful periods with cramps** | 40 | 64.5 | 11 | 28.9 | |
| | P6 | **Menorrhagia (Heavy Bleeding)** | 21 | 33.9 | 9 | 23.7 | *Derived from P32* |
| **Pathological sequelae (reproductive anomalies)** | P2 | **PCOS/PCOD** | 19 | 30.6 | 0 | 0 | |
| | P7 | **Anovulation** | 1 | 1.6 | 0 | 0 | |
| | P10 | **Tubal Blockage** | 1 | 1.6 | 0 | 0 | *Same as P16* |
| | P11 | **Secretory/Proliferative/thickened Endometrium/Endometrium hyperplasia/Endometrial Cysts** | 4 | 6.5 | 0 | 0 | |
| | P12 | **Teratoma (Ovarian Cyst)** | 1 | 1.6 | 0 | 0 | |
| | P13 | **Abortions/Medical termination of pregnancy** | 16 | 25.8 | 0 | 0 | |
| | P14 | **Endometriosis** | 2 | 3.2 | 0 | 0 | |
| | P16 | **Fallopian tubes or uterine blockage suggested by Hysterosalpingogram (X-ray of uterus/tubes)** | 5 | 8.1 | 0 | 0 | |
| **Genital infection associated** | P8 | **Vaginal discharge/leukorrhea** | 1 | 1.6 | 0 | 0 | *Same as P21* |

| | | | | | | |
|---|---|---|---|---|---|---|
| **symptoms/confirmed diagnosis of infections** | P9 | **Thrombocytopenia** | 1 | 1.6 | 0 | 0 |
| | P15 | **Dyspareunia (pain during intercourse)** | 7 | 11.3 | 0 | 0 |
| | P17 | **Cervicitis (Cervical inflammation)** | 2 | 3.2 | 0 | 0 |
| | P18 | **TB Positivity PPD/Quantiferon TB test/TB PCR test/TB culture test done or not ?** | 2 | 3.2 | 0 | 0 |
| | P19 | **Confirmed/Symptoms of UTI (painful micturition)** | 2 | 3.2 | 0 | 0 |
| | P20 | **Pelvic Inflammatory Disorder** | 16 | 25.8 | 0 | 0 |
| | P21 | **Vaginitis/symptoms of infections (Itching, burning, discharge)** | 11 | 17.7 | 0 | 0 |
| **Female Treatment** | P22 | **Treatment with Clomid and/or Letrozole alone** | 24 | 38.7 | 0 | 0 |
| | P23 | **Treatment with Estrogen Priming followed by Letrozole and/or Clomid** | 25 | 40.3 | 0 | 0 |
| | P24 | **Treatment with Letrozole/Clomid priming followed by Progesterone** | 1 | 1.6 | 0 | 0 |
| | P25 | **Treatment with Estrogen Alone** | 1 | 1.6 | 0 | 0 |
| | P26 | **Treatment with Progesterone Alone** | 1 | 1.6 | 0 | 0 |
| **Physical parameters** | P35 | **Do you exercise or workout?** | 22 | 35.5 | 15 | 39.5 |
| **Male reproductive/ semen/sperm anomalies** | P36 | **Was the semen abnormal?** | 23 | 37.1 | 0 | 0 |
| | P37 | **Oligospermia (Low Sperm Count)** | 13 | 21 | 0 | 0 |
| | P38 | **Low Sperm DNA Integrity** | 2 | 3.2 | 0 | 0 |
| | P39 | **Asthenozoospermia (AS) or Low motility Sperm** | 14 | 22.6 | 0 | 0 |
| | P40 | **High TZI (Teratozoospermia index) levels** | 9 | 14.5 | 0 | 0 |
| | P41 | **Normospermia** | 8 | 12.9 | 0 | 0 |
| | P42 | **Pyospermia [Presence of pus cells]** | 5 | 8.1 | 0 | 0 |
| | P43 | **Hypospermia [Low volume of semen]** | 2 | 3.2 | 0 | 0 |
| | P44 | **Hyperviscosity** | 3 | 4.8 | 0 | 0 |
| **Male Treatment** | P45 | **Treated for TB with Macoz (Rifampicin and Isoniazid)** | 1 | 1.6 | 0 | 0 |

**B.**

| | | P33: Details of menstrual flow[a] |
|---|---|---|
| **Total frequency of infertility group, N1 = 62** | **Frequency of patients with heavy menstrual bleeding (F1)** | 21 |
| | **Infertile % with heavy menstrual bleeding (F1*100/N1)** | 33.9 |
| | **Frequency of patients with light menstrual bleeding (F2)** | 3 |
| | **Infertile % with light menstrual bleeding (F2*100/N1)** | 4.8 |
| | **Frequency of patients with moderate menstrual bleeding (F3)** | 38 |
| | **Infertile % with moderate menstrual bleeding (F3*100/N1)** | 61.3 |
| **Total frequency of healthy control group, N2 = 38** | **Frequency of healthy subjects with heavy menstrual bleeding (F4)** | 9 |
| | **Healthy % with heavy menstrual bleeding (F4*100/N2)** | 23.7 |
| | **Frequency of healthy subjects with light menstrual bleeding (F5)** | 0 |
| | **Healthy % with light menstrual bleeding (F5*100/N2)** | 0 |
| | **Frequency of healthy subjects with moderate menstrual bleeding (F6)** | 29 |
| | **Healthy % with moderate menstrual bleeding (F6*100/N2)** | 76.3 |
| a: Menstrual flow data is sub-categorized into three categories- Light, moderate and heavy | | |

**C.**

| | | P34: Physical parameter (BMI)[b] |
|---|---|---|
| **Total frequency of infertility group, N1 = 62** | **Frequency of patients with Normal BMI (F1)** | 19 |
| | **Infertile % with normal BMI (F1*100/N1)** | 30.6 |
| | **Frequency of patients with high BMI (F2)** | 10 |
| | **Infertile % with high BMI (F2*100/N1)** | 16.1 |
| | **Frequency of patients with low BMI (F3)** | 7 |
| | **Infertile % with low BMI (F3*100/N1)** | 11.3 |
| | **Frequency of patients with no BMI data (F4)** | 26 |
| | **Infertile % with no BMI data (F4*100/N1)** | 41.9 |
| **Total frequency of healthy control group, N2 = 38** | **Frequency of healthy with Normal BMI (F5)** | 16 |
| | **Healthy % with normal BMI (F5*100/N2)** | 42.1 |
| | **Frequency of healthy with high BMI (F6)** | 6 |
| | **Healthy % with high BMI (F6*100/N2)** | 15.8 |
| | **Frequency of healthy with low BMI (F7)** | 0 |
| | **Healthy % with low BMI (F7*100/N2)** | 0 |
| | **Frequency of healthy with no BMI data (F8)** | 16 |
| | **Healthy % with no BMI data (F8*100/N2)** | 42.1 |
| b: BMI data is sub-categorized into three categories- Low, normal and high | | |

**D.**

| | | Physiological parameters (female reproductive hormone profiles) | | | |
|---|---|---|---|---|---|
| | | **P27** | **P28** | **P29** | **P30** |
| | | **Follicle stimulating hormone (FSH)[c]** | **Luteinising hormone (LH)[c]** | **Prolactin hormone[d]** | **Anti-Mullerian Hormone (AMH)[d]** |
| **Total frequency of infertility group, N1 = 62** | **Frequency of patients with high hormone levels (F1)** | 3 | 1 | 2 | 6 |
| | **Infertile % with high hormone levels (F1*100/N1)** | 4.8 | 1.6 | 3.2 | 9.7 |
| | **Frequency of patients with low hormone levels (F2)** | 2 | 2 | 0 | 0 |
| | **Infertile % with low hormone levels (F2*100/N1)** | 3.2 | 3.2 | 0 | 0 |
| | **Frequency of patients with normal hormone levels (F3)** | 31 | 5 | 9 | 8 |
| | **Infertile % with normal hormone levels (F3*100/N1)** | 50 | 8.1 | 14.5 | 12.9 |
| | **Frequency of patients with no hormone data (F4)** | 26 | 54 | 51 | 48 |
| | **Infertile % with no hormone data (F4*100/N1)** | 41.9 | 87.1 | 82.3 | 77.4 |
| | **Healthy (N2 = 38)** | No data | No data | No data | No data |

c: FSH and LH levels are sub-categorized into three categories- Low, high and normal
d: Prolactin and AMH are sub-categorized only into two categories- High and normal (hence, low frequency is listed as 0)
